# Variable pharmacokinetics of coagulation factor VIII in the perioperative setting complicates personalisation of treatment in patients with haemophilia A

**DOI:** 10.1101/2025.02.07.25321866

**Authors:** Alexander Janssen, Iris van Moort, Frank C. Bennis, Marjon H. Cnossen, Ron A.A. Mathôt

## Abstract

Pharmacokinetic (PK)-guided dosing of factor concentrates in patients with haemophilia A is generally recommended for the optimisation of prophylactic treatment. PK-guided dosing can also be useful in the perioperative setting, where guidelines advise to keep factor VIII (FVIII) activity levels within tight target ranges to prevent bleeding. Previous studies suggest changes in FVIII PK following medical procedures as well as potential time-dependent effects, meaning that population PK models specific to the perioperative setting are required. In this study, we use data from haemophilia A patients collected during the prophylactic and perioperative setting to identify covariates that explain changes in FVIII PK. Additionally, we use machine learning methods to find potential time-dependent effects on FVIII clearance. Perioperative FVIII clearance was generally lower compared to the prophylactic setting. Covariates related to the complexity of the medical procedures were correlated to larger decreases in clearance. Importantly, subjects undergoing more complex procedures also depicted potentially relevant time-dependent effects on clearance. These effects could be highly variable between subjects. Directly using PK parameters obtained from the prophylactic setting resulted in relatively high mean absolute percentage error (MAPE) at 26.3%, while the perioperative model including time-dependent effects depicted markedly reduced error (10.3%). The presence of high variability between subjects and potential time-dependent effects complicates the selection of optimal dosing regimens before the start of medical procedures. Our method can be used to optimise treatment in real-time, but close monitoring of FVIII levels will likely remain necessary.

## Introduction

Clinical guidelines advise pharmacokinetic (PK-)guided dosing of factor VIII (FVIII) concentrates to personalise prophylactic treatment for patients with haemophilia A [1, 2]. In this context, population PK models are used to produce individual estimates of PK parameters based on measured FVIII levels. These parameters can be used to simulate expected FVIII exposure based on different dosing regimens, and enables the selection of an optimal treatment strategy that meets pre-specified target levels. Several population PK models have already been developed for this purpose, of which the majority are specific to a single brand of FVIII concentrate [3]. Another clinical scenario where PK-guided dosing might be useful is following medical procedures. Patients with haemophilia face particular risks in the perioperative setting [2]. One of the main risks is a higher chance of bleeding complications, which is exacerbated in major versus minor procedures [2, 4]. Unfortunately, data on optimal FVIII peak and trough levels to maintain after these procedures is not conclusive [5]. Clinical guidelines, primarily based on expert opinion, suggest to maintain high levels of FVIII during the procedure and to gradually decrease target levels over the subsequent days [5, 6]. Generally, body weight-based dosing via intermittent bolus injections or continuous infusions is advised. However, several studies have shown that patients are regularly under- and overdosed when performing weight-based dosing in the perioperative setting [7–9]. The OPTI-CLOT randomized controlled trial (RCT) by van Moort et al. directly compared weight-based to PK-guided dosing and found that the latter resulted in improved FVIII target attainment (68% of measurements within the target range versus 37% for weight-based dosing) [8].

Despite these findings, only a single population PK model for the perioperative setting has been described in the literature [10]. This retrospective study by Hazendonk et al. describes a model for moderate and severe haemophilia A patients who underwent elective, minor, and major medical procedures in The Netherlands [10]. The covariate analysis indicated decreased clearance in older individuals and those undergoing higher-risk procedures. Importantly, residual between-subject variability was still relatively large (>25% coefficient of variation) and potentially important covariates such as von Willebrand factor (VWF) were missing from the model. Furthermore, the study could not determine differences in the PK parameters compared to the prophylactic setting as only perioperative FVIII levels were collected. Finally, no typical time-dependent effect could be detected, although their presence has been suggested by previous studies [11–13]. Later studies extending and validating the model on paediatric and external patient populations indicated that model adjustments were required [14, 15].

In this study, we aim to develop a population PK model that resolves these issues. We use the data collected during the OPTI-CLOT RCT [8]. In this trial, each participant received a PK profile before their medical procedure. Additionally, frequent FVIII monitoring was performed before and after each medical procedure, facilitating the analysis of time-dependent effects. This data is thus well-suited for the development of a model that describes the PK of FVIII during the prophylactic setting (based on individual PK profiles) as well as during the perioperative setting. Our approach is as follows: first, we apply a recently described machine learning (ML)-based method for covariate analysis to identify influential covariates and their relationship with FVIII clearance and volume of distribution [16]. Based on this information, we develop a population PK model on the PK profile data and produce individual PK parameter estimates specific to the prophylactic setting. We then adapt these parameters to data from the perioperative setting using the covariates. Finally, we use Gaussian Processes, another ML method, to identify and correct for any time-dependent effects on FVIII clearance on an individual basis.

## Methods

### Data

Data from 69 moderate and severe haemophilia patients were collected during the OPTI-CLOT RCT [8]. In this study, the FVIII consumption of body weight-based and PK-guided dosing was compared during elective, low, and medium risk medical procedures. Patients were stratified according to the severity of the procedure and the mode of FVIII administration (bolus or continuous infusion). First, a PK profile was constructed for each enrolled patient. In this context, FVIII plasma samples were collected at approximately 4, 24, and 48 hours after a test dose. Only PK profile data was available for three patients as their medical procedure was cancelled. The 66 remaining patients underwent a medical procedure within a period of one year after the PK profile assessment. In the perioperative setting, patients received bolus and/or continuous infusions of standard half- life FVIII concentrate in order to meet pre-specified FVIII target levels according to national clinical guidelines [6].

Several blood samples were collected for each patient (both before and after the procedure). FVIII levels were monitored at 24 hour intervals up until a maximum of 14 days post-procedure or until the patient was discharged. Most patients (n = 47) had a follow-up time greater than 48 hours. A total of 228 measurements (median 3 per patient) were available at the time of the PK profiles and 568 measurements (median 9 per patient) were collected around the medical procedures. All FVIII measurements were performed using the one-stage assay. Potentially relevant patient characteristics included patient body weight, height, age, blood group, pre-assessed surgical risk score (low or medium), haemophilia severity (moderate or severe), treatment centre, expected and actual duration of the procedure, expected and actual blood loss during the procedure (mild or moderate), NaCl administration (mL), brand of FVIII concentrate, von Willebrand factor antigen (VWF:Ag), activity (VWF:act), and pro-peptide (VWF:pp) levels, and finally the occurrence of bleeds after the procedure. Additional covariates calculated from the data were BMI and fat-free mass (FFM; using Al Sallami’s equation [17]).

### Imputation of missing values

Some of the covariates contained missing values. Most prominently, VWF antigen (VWF:Ag) levels were missing for a majority of the patients at the time of their PK profile. A recently described generative model was used to set prior distributions over VWF:Ag, and maximum a posteriori (MAP) estimation was performed during model development to obtain individual estimates [18]. For other covariates (body weight and height), a multiple imputation strategy was employed in order to evaluate the sensitivity of the analysis to the imputation procedure. Ten replicates of imputed data sets were produced based on samples from the generative model. More information on the applied strategy can be found in Appendix B.

### Population PK model development

A population PK model was developed to transform individual PK parameter estimates obtained from the prophylactic setting to the perioperative setting. First, a non-linear mixed effect (NLME) model was developed based on the 69 available PK profiles. Covariate selection was informed by using a recently described ML method that learns the covariate implementation directly from data.SHapley Additive exPlanations (SHAP), and explainable AI method, is then applied to visualize the learned effects [16]. Based on these visualizations, covariates were selected for inclusion using a full-model based approach, and retained if their removal resulted in a significant increase in objective function value (p < 0.01, χ^2^ critical value: 6.635), there were expected pharmacological/causal relationships (based on expert assessment), or when removal resulted in a relevant increase in prediction bias. Prediction error was evaluated using the mean absolute percentage error (MAPE), root mean squared error (RMSE), and coefficient of determination (R^2^). The above procedure was repeated to produce the perioperative model. Differently however, the SHAP analysis was performed on the difference between PK parameter estimates from the final prophylactic model and individual perioperative PK parameters estimated using a base NLME model. The results from this analysis thus identifies potentially relevant covariates that explain the difference in the PK parameters between the two settings. Covariates were again included using a full-model approach, and models for each of the ten imputation replicates were critiqued in the same way as for the prophylactic model to produce the final model. A complete step-by-step guide through model development and decisions is available in appendix B.

### Learning time-dependent effects

Previous results indicate that FVIII PK might vary in the days following the medical procedure. To test this hypothesis, time-dependent effects on FVIII clearance were added to the perioperative model. Gaussian Process models were used to learn time-dependent effects on an individual basis. Gaussian Processes are flexible models representing an extension of multivariate normal distributions in continuous time (i.e. in infinite dimensions), and are well suited for this problem [19, 20]. For each subject, we can use these models to obtain a posterior distribution over the change in FVIII clearance at each time point based on discrepancies (if any) between FVIII measurements and model predictions. Time-dependent effects were added for subjects with a follow-up time greater than 48 hours (n = 47). Details on the exact implementation can be found in appendix B. Patients were grouped according to the maximal percentage change in clearance during follow-up. A change in FVIII clearance greater than 15% was deemed potentially relevant. We then performed statistical tests to evaluate which covariates were significantly correlated to the subjects presenting potentially relevant effects. For continuous covariates, two-tailed t-tests were performed. For categorical variables, chi-squared tests were performed. A critical value of 0.05 was used to determine significance.

## Results

### Patient characteristics

Information on patient characteristics during the prophylactic and perioperative setting are shown in table 1. At the time of the PK profiles, patient height was missing for the three patients who did not undergo a medical procedure. Additionally, VWF:Ag measurements were missing during the prophylactic setting for a large proportion of patients (65%). Contrastingly, multiple VWF measurements were performed in the perioperative setting. In this setting, only nine patients presented with missing information. A correlation matrix depicting covariate relationships at the time of the surgical procedures is shown in supplementary figure 1.

**Table 1.**
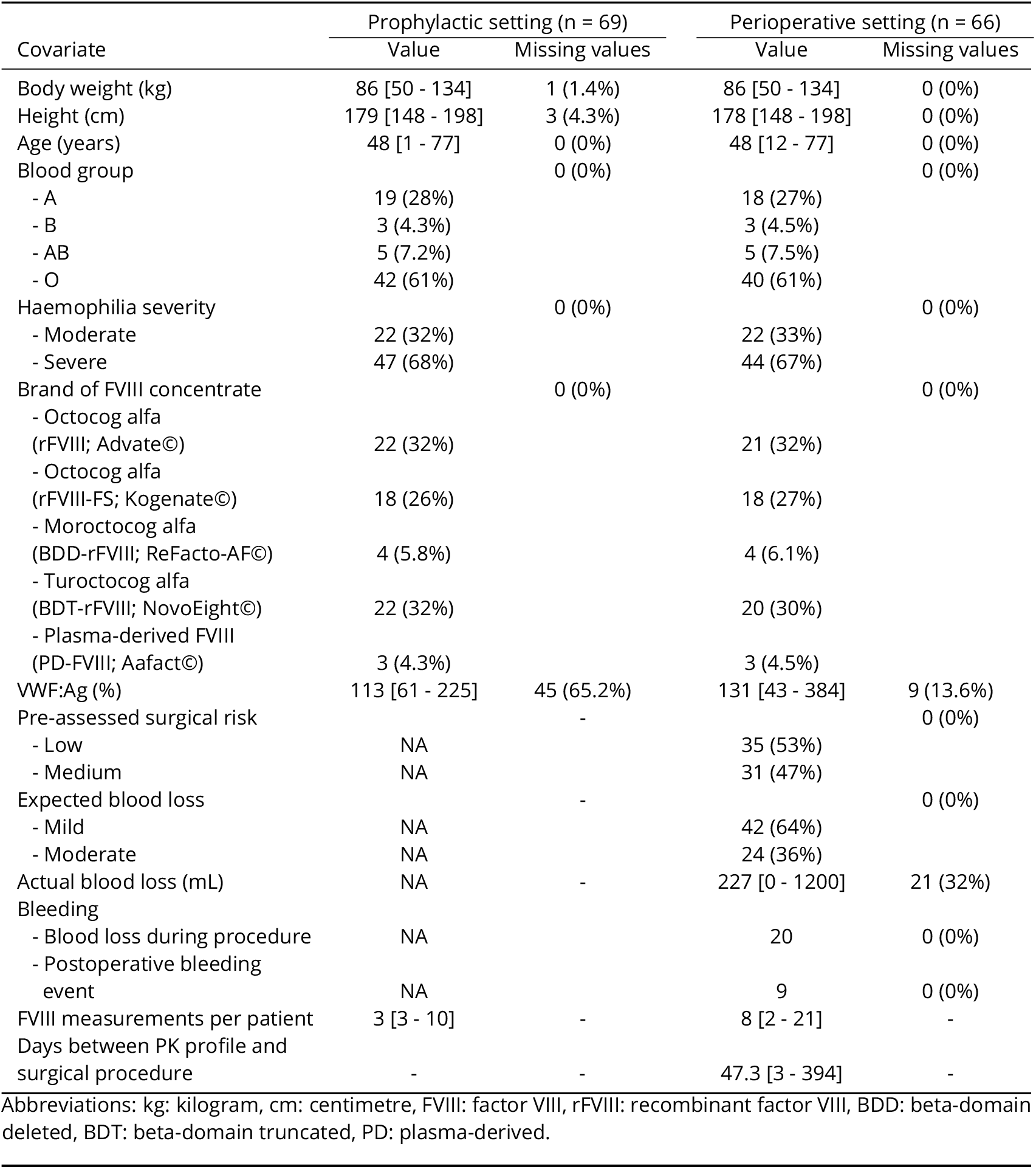
Patient characteristics.

### Differences in PK parameters

An overview of the covariate effects included in the final prophylactic and perioperative population PK models is shown in figure 1. Details on the covariate selection process is available in appendix B. In the prophylactic model, the effect of FFM on clearance and volume of distribution was best explained using a linear effect (coefficient of 1). After FFM, VWF:Ag was the most important parameter explaining variability in FVIII clearance (linear coefficient of 0.39, 95% CI [0.30, 0.48]). Clearance was slightly lower in individuals who received rFVIII-FS (−21%, 95% CI [−33%, −9.2%]). Similarly, individuals who received BDD-rFVIII or those treated in centre 2 had overall lower FVIII levels compared to other patients (−20%, 95% CI [−35%, −6%] and −15% [−29%, −2%], respectively). Coefficient of variation for the between-subject variability of clearance (18%) and volume of distribution (12%) were relatively low. MAPE of predictions based on individual estimates of the PK parameters was 19.1% (RMSE = 7.90 IU/dL, R^2^ = 0.97).

**Figure 1.**
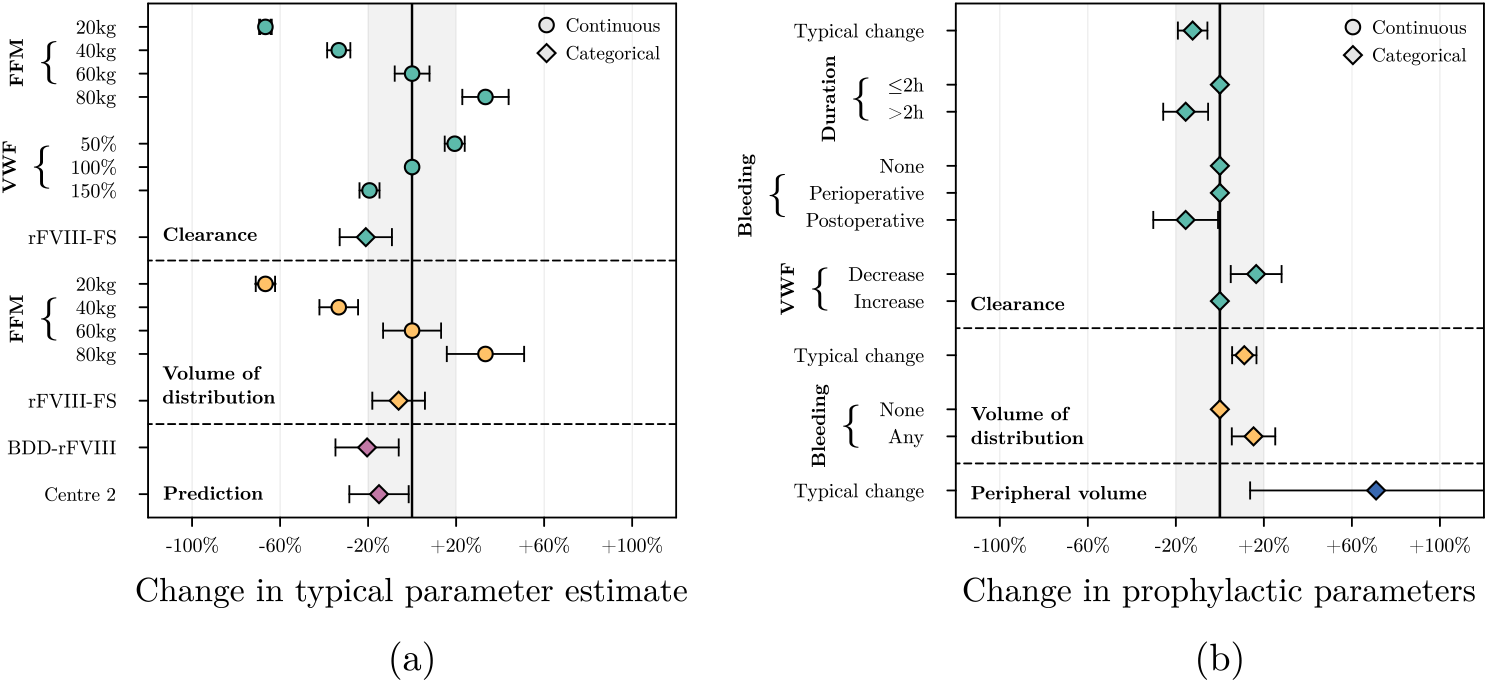
Forest plot depicting covariate effects in the population PK models. Parameter estimates along with 95% confidence intervals (CI) are shown for the models fit on data from the prophylactic (a) and the perioperative (b) setting. Circles (continuous variables) and diamonds (categorical variables) indicate mean parameter value across training replicates and error bars represent the CI. Coloured region indicates covariate effects resulting in <20% change in the parameters. Abbreviations: FFM = fat-free mass, VWF = von Willebrand factor, rFVIII-FS = recombinant factor VIII formulated with sucrose, BDD-rFVIII = beta-domain deleted recombinant factor VIII.

Compared to the prophylactic setting, FVIII clearance was lower in all patients during the entire perioperative window (−13%, 95% CI [−19%, −6%]). Similarly, FVIII volume of distribution was higher, both in the central (+11%, 95% CI [+6.6%, +15%]) and peripheral compartments (+73%, 95% CI [+12% +234%]). Lower FVIII clearance was seen in patients undergoing procedures with a duration longer than two hours (−16%, 95% CI [−32%, +1%]) and those who had a postoperative bleed (−16%, 95% CI [−26%, −6%]). Patients with reported blood loss during the procedure or with a postoperative bleed had roughly 15% higher volume of distribution (95% CI [+6%, +24%]) compared to patients without bleeding. Next, higher clearance was seen for patients with a decrease in perioperative VWF:Ag levels relative to the estimated prophylactic levels (+16%, 95% CI [+5%, +27%]). According to the prophylactic model, subjects treated in centre 2 had relatively lower FVIII levels compared to the other treatment centres. This effect was re-estimated in the perioperative model and found to be unimportant. The effect was consequently removed in the final model. In contrast, the effect of BDD-rFVIII on observed FVIII activity levels (−21%, 95% CI [−39% −2%]) was very similar and thus retained in the model.

Prediction error (MAPE) was 24.9% (RMSE = 26.4 IU/dL, R^2^ = 0.63) when using the individual PK parameters estimated from the prophylactic data to make predictions in the perioperative setting. After adjusting for the aforementioned covariate effects, prediction error improved to 20.9% (RMSE = 21.9 IU/dL, R^2^ = 0.74). Coefficient of variation for the between-subject variability on clearance (18%) and volume of distribution (11%) were again relatively low. When estimating individual PK parameter estimates using the perioperative model, MAPE further improved to 13.9% (RMSE = 16.9 IU/dL, R^2^ = 0.85).

### Time-dependent effects on FVIII clearance

Time-dependent effects on FVIII clearance were added to the final perioperative model to investigate whether any relevant changes could be observed. The addition of time-dependent effects led to a further improvement of the MAPE from 13.9% to 10.7% (RMSE = 14.0 IU/dL, R^2^ = 0.90). The addition of time-dependent effects on clearance seemed to mainly result in improved predictive performance in the first two days after the procedure (judging from the lower variability of residual error; see supplementary figure 2). Importantly, the model did not indicate that FVIII clearance returned to prophylactic levels during follow-up. An overview of the predictions with and without time-dependent effects for each individual is provided in supplementary figure 3.

We found that patients could be divided into two groups: (1) those with potentially relevant changes (>15% maximal change, n = 25) and (2) those with minor changes (0-15% change, n = 22), see figure 2. Variability in the effect on FVIII clearance for those with potentially relevant effects was relatively large, with the maximal effect ranging from a 15 to 100% change (figure 2a). When looking specifically at the prediction error for subjects in this group, MAPE improved from 17.0% (RMSE = 14.1 IU/dL, R^2^ = 0.80) to 11.6% (RMSE = 10.7 IU/dL, R^2^ = 0.89) after taking into account time-dependent effects. An example of the prediction for a subject with relevant time-dependent effects is shown in figure 3.

**Figure 2.**
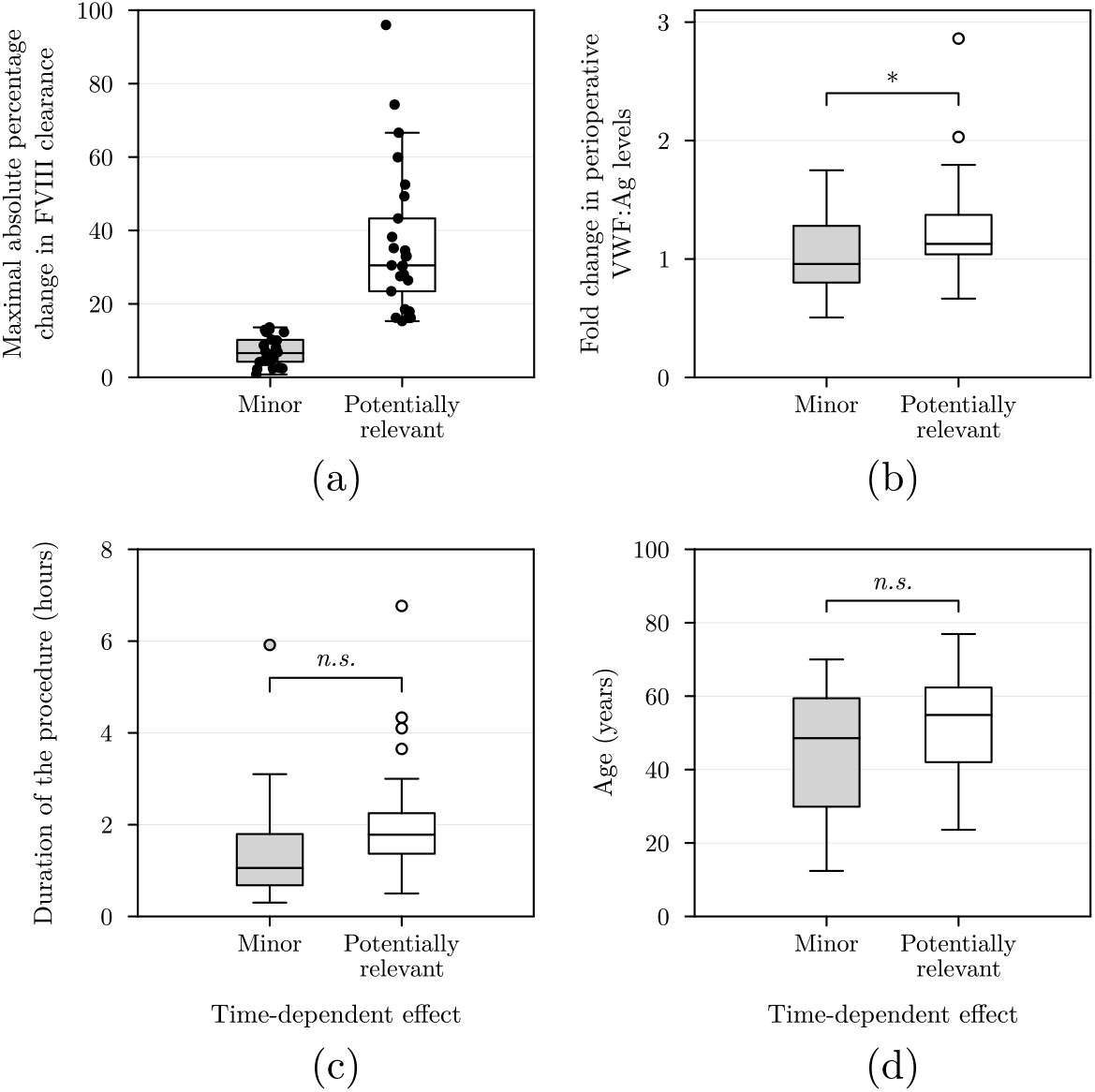
Time-dependent effects and relevant covariates. Here we compare subjects with minor or potentially relevant time-dependent effects. In (a), we show the maximal change in FVIII clearance from the time-dependent effect for each of the subjects. In the other figures, we compare differences between subjects with minor or potentially relevant time-dependent effects based on the change in perioperative VWF:Ag levels (b), duration of the medical procedure (c), or age (d). Asterisk (*) indicates statistical significance (*p* < 0.05).

**Figure 3.**
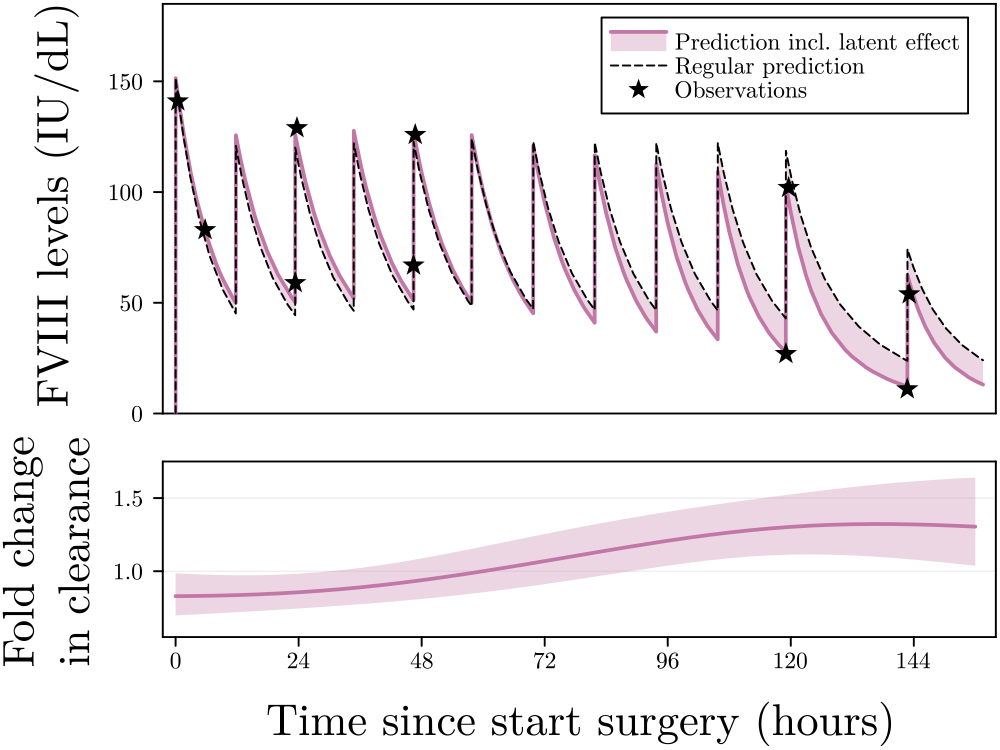
Example of the prediction for a subject with relevant time-dependent effects. In the top panel, observed FVIII activity levels (stars) and model predictions with (coloured line) and without (dashed line) time-dependent effects are shown. Filled area depicts the difference between the two predictions. In the bottom panel, the time-dependent effect on FVIII clearance (mean + 95% credible interval) is shown. The presented change in clearance is with respect to the typical perioperative clearance estimate for this individual. We can see that FVIII clearance seems to be relatively lower in the first two days after surgery compared to later time points. Without this correction, FVIII levels at later time points are over-estimated, potentially resulting in overconfident treatment.

Next, we looked at covariates that could identify subjects who presented potentially relevant effects. We found that the change in perioperative VWF:Ag relative to the estimated prophylactic levels was significantly different between the two groups (t = −2.02, p < 0.05; figure 2b). Moreover, subjects who had an increase in perioperative VWF:Ag levels were significantly more likely to depict relevant time-dependent effects (χ^2^ = 4.62, p < 0.05). Patient age, duration of the medical procedure, and surgical risk score were correlated to the presence of time-dependent effects but did not reach statistical significance (figure 2c & d).

## Discussion

The aim of this study was to identify covariates that were predictive of the differences in FVIII PK between the prophylactic and perioperative setting. To this end, we developed a population PK model that adapts individual PK parameter estimates from the prophylactic setting to the perioperative setting. Here, we found that FVIII clearance was generally lower following medical procedures. Subjects undergoing longer surgical procedures (>2 hours) as well as those who presented post- operative bleeding depicted additional reductions in clearance. Patients for whom VWF:Ag levels decreased with respect to the prophylactic setting had higher clearance. Next, FVIII volume of distribution was slightly higher in the perioperative setting, and blood loss (both during procedures and as a result of postoperative bleeding) resulted in additional increases. We found that patients who received BDD-rFVIII had lower FVIII levels, which matches earlier findings related to assay discrepancies [21]. The addition of covariates slightly improved prediction accuracy (MAPE 24.9% to 20.9%) compared to when prophylactic PK parameters were used. Our analysis also indicated the presence of potentially relevant time-dependent effects on FVIII clearance in 25 individuals (further reducing MAPE to 10.7%). Patients with larger perioperative changes in VWF:Ag were significantly more likely to present relevant time-dependent changes in FVIII clearance. Importantly, in none of the patients did the PK parameters return to levels from the prophylactic setting. In contrast to a earlier population PK model specific to the perioperative setting, we did identify time-dependent effects on FVIII PK. We dedicate this finding to the learning of individual level effects (through the use of Gaussian Processes) instead of estimating population level effects. It is likely that previous studies were unable to find shared effects due to the high variability in the time-dependent effect. In addition, we found that roughly half of subjects depicted no time-dependent effects, further complicating the learning of a shared effect. We identified several factors that were related to the subject presenting potentially relevant time-dependent effects. It is likely that the observed correlations to changes in perioperative VWF:Ag levels, patient age, and the duration of surgery are confounding effects. Increases in perioperative VWF:Ag levels could be related to vascular damage and stress responses following the surgical procedure [22, 23]. Patients with relevant effects also tended to be older patients, who are more likely to undergo more complex procedures. Similarly, surgical procedures of longer duration are also likely more complex or have faced complications. It is possible that haemostatic challenges sustained during complex procedures result in deviations in FVIII clearance in the days following the procedure. The variability in the effect might be related to the individual stress response and degree of damage sustained during medical procedures for each patient. Similarly, the covariates identified in the perioperative population PK model might also be related to downstream effects of the medical procedure. For example, patients undergoing longer procedures and those with postoperative bleeds had lower FVIII clearance, which might again be related to the complexity of the procedure. Similarly, FVIII volume of distribution was higher when the patient suffered blood loss, which might be related to the leakage of FVIII from the damaged site or the injection of plasma/saline solution. Although we can find reasonable rationales for including these covariates, their effects were relatively small. It is possible that their effects were also temporary, such that adding their effects on the PK parameters for the entire perioperative time window results in underestimation of their importance. For example, the consequences of bleeding for FVIII volume of distribution of course occurs only after the bleed in question has actually happened. In the current implementation of the model, these covariate effects affected PK from the start of the procedure. Future work should investigate if time-dependent estimation of covariate effects improves predictions. Findings from our study indicate important consequences for PK-guided dosing of factor concentrates in the perioperative setting. First, individual estimates of the PK parameters obtained from the prophylactic setting were not sufficient to adequately predict perioperative FVIII levels. As a consequence, these parameters might give biased predictions of FVIII levels when selecting the initial treatment schedule before medical procedures. Next, information on the covariates important in the perioperative model will likely only become available in the days following the procedure. This complicates real-time optimisation of treatment. Finally, the presence of potentially relevant time-dependent effects on FVIII clearance complicates the prediction of future FVIII levels. Since these effects were highly specific to each individual, close monitoring of FVIII levels will likely still be required during the perioperative setting. This process will likely be time-intensive, and most treatment centres might lack the expertise to facilitate such analyses. More research is thus required to provide user-friendly tools to facilitate PK-guided dosing in the perioperative setting. There also were some limitations to our study. First, time-dependent effects were estimated in a post hoc manner, e.g. after including the covariate effects. As mentioned before, the time-dependent effects might be related to the effects of the included covariates, and thus re-estimation of the covariate effects might be preferred. Next, covariate selection for the perioperative model was complicated by subtle differences in patient populations between treatment centres. As a result, some factor concentrates or treatment centres depicted relevant correlations to changes in the PK parameters or the presence of time-dependent effects. Selection of the appropriate causal covariate in this setting can be complicated. Finally, the OPTI-CLOT RCT mainly included adult patients undergoing minor, elective or medium risk medical procedures. The covariate effects in a paediatric population, as well as the changes in the PK parameters following major surgery cannot be induced from our model. It is likely that relevant time-dependent effects are also present following major surgery, but additional research is required in order to determine if and how these effects would differ from those found in our study. In conclusion, we describe a perioperative population PK model that can be used to adjust FVIII PK parameters from the prophylactic setting to the perioperative setting. We also found that roughly half of patients faced relevant time-dependent changes in FVIII clearance. Our analysis is indicative of the difficulty of predicting perioperative FVIII levels, and shows that PK-guided dosing in this setting is complicated by additional subject-specific variability in PK. Frequent monitoring of FVIII following medical procedures might thus still be required. Future research should focus on identifying additional sources explaining the remaining inter-individual variability and time-dependent effects in order to support PK-guided dosing of factor concentrates in the perioperative setting.

## Data Availability

All model code is made available at https://github.com/JanssenaPerioperativeFVIII.

## Acknowledgments

SYMPHONY consortium: The SYMPHONY consortium which aims to orchestrate personalized treatment in patients with bleeding disorders, is a unique collaboration between patients, health care professionals, and translational and fundamental researchers specialized in inherited bleeding disorders, as well as experts from multiple disciplines. It aims to identify best treatment choice for each individual based on bleeding phenotype. To achieve this goal, workpackages have been organized according to 3 themes e.g. Diagnostics (workpackages 3 and 4); Treatment (workpackages 5-9) and Fundamental Research (workpackages 10–12). This research receives funding from the Netherlands Organisation for Scientific Research (NWO) in the framework of the NWA-ORC Call grant agreement NWA.1160.18.038. Principal investigator: Dr. M.H. Cnossen. Project coordinator: Dr. S.H. Reitsma.

Beneficiaries of the SYMPHONY consortium: Erasmus University Medical Center-Sophia Children’s Hospital, project leadership and coordination; Sanquin Diagnostics; Sanquin Research; Amsterdam University Medical Centers; University Medical Center Groningen; University Medical Center Utrecht; Leiden University Medical Center; Radboud University Medical Center; Netherlands Society of Hemophilia Patients (NVHP); Netherlands Society for Thrombosis and Hemostasis (NVTH); Bayer B.V., CSL Behring B.V., Swedish Orphan Biovitrum (Belgium) BVBA/SPRL. Additional beneficiaries, not included in the SYMPHONY consortium, currently funding parallel projects are: Novonordisk (OPTI-CLOT TARGET), Roche (Partitura), Stichting Haemophilia (patient-reported outcomes project).

OPTI-CLOT study group: OPTI-CLOT/To WiN study group aims to implement personalized treatment by pharmacometric-guided dosing of factor concentrates, desmopressin and nonfactor therapies in patients with bleeding disorders.

OPTI-CLOT/To WiN Steering Committee, the Netherlands: M.H. Cnossen (principal Investigator & chair OPTI-CLOT-To WiN) and R.A.A. Mathôt (coinvestigator). F.W.G. Leebeek, Rotterdam;, M. Coppens K. Fijnvandraat, Amsterdam; K. Meijer, Groningen, S.E.M. Schols, Nijmegen; H.C.J. Eikenboom, Leiden; R.E.G. Schutgens, Utrecht; F. Heubel-Moenen, Maastricht; L. Nieuwenhuizen, Veldhoven; P. Ypma, The Hague; M.H.E. Driessens, Nijkerk.

Trial bureau: I. van Vliet, Rotterdam.

Local collaborators the Netherlands: M.J.H.A. Kruip, S. Polinder, Rotterdam; P. Brons,Nijmegen;

F.J.M. van der Meer, Leiden; K. Fischer, K. van Galen, Utrecht.

Principal investigators and local collaborators in the UK: P. W. Collins, Cardiff; M. Mathias, P. Chowdary, London; D. Keeling, Oxford.

OPTI-CLOT-To WiN, DAVID and SYMPHONY PhDs: PhDs: J. Lock, H.C.A.M. Hazendonk, T. Preijers,

N.C.B. de Jager, L. Schutte, L.H. Bukkems.

PhDs ongoing: M.C.H.J. Goedhart, J.M. Heijdra, L. Romano, W. Al Arashi, M.E. Cloesmeijer, A. Janssen, S.F. Koopman, C. Mussert.

This preprint was typeset using the LaPreprint template (https://github.com/roaldarbol/lapreprint) by Mikkel Roald-Arbøl.

## Author contributions

Conceptualization and design: AJ, Data acquisition: IM, Methodology, data analysis and interpretation: AJ, Original draft: AJ, Writing - review & editing: AJ, IM, MC, RM, Final approval of the version to be published: AJ, IM, MC, RM.

