## Supplementary data for "Variable pharmacokinetics of coagulation factor VIII in the perioperative setting complicates personalisation of treatment in patients with haemophilia A"

### APPENDIX

#### 8.A SUPPLEMENTARY FIGURES

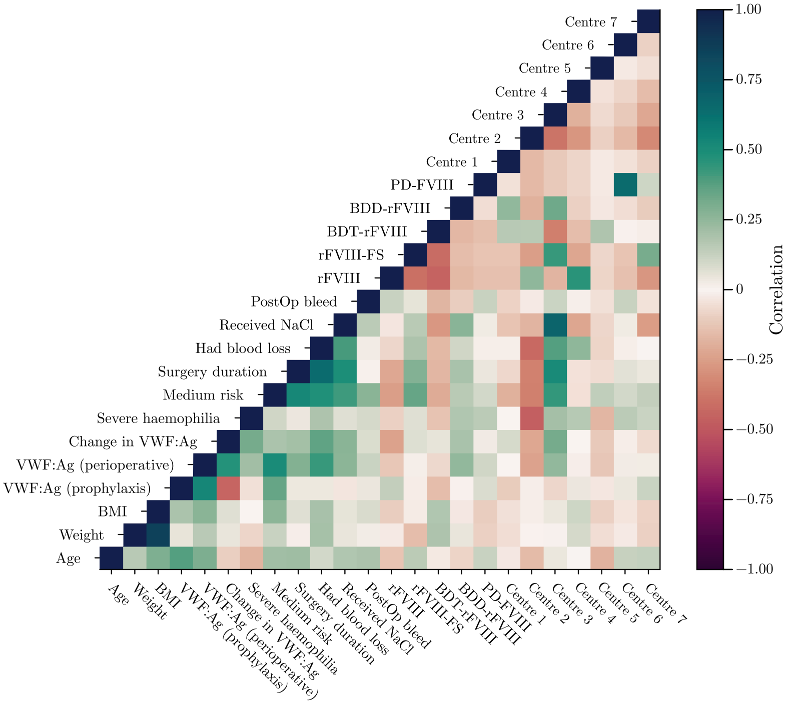


Figure 8.A.1: Correlation between relevant covariates at the time of the surgical procedures. Here, the correlation between relevant covariates and the change in individual PK parameter estimates with respect to the prophylactic setting are shown. Importantly we can observe relevant correlations between different treatment centres or different rFVIII concentrates and variables related to the surgical procedures (e.g. risk score, perioperative change in VWF:Ag, blood loss during surgery, etc.).

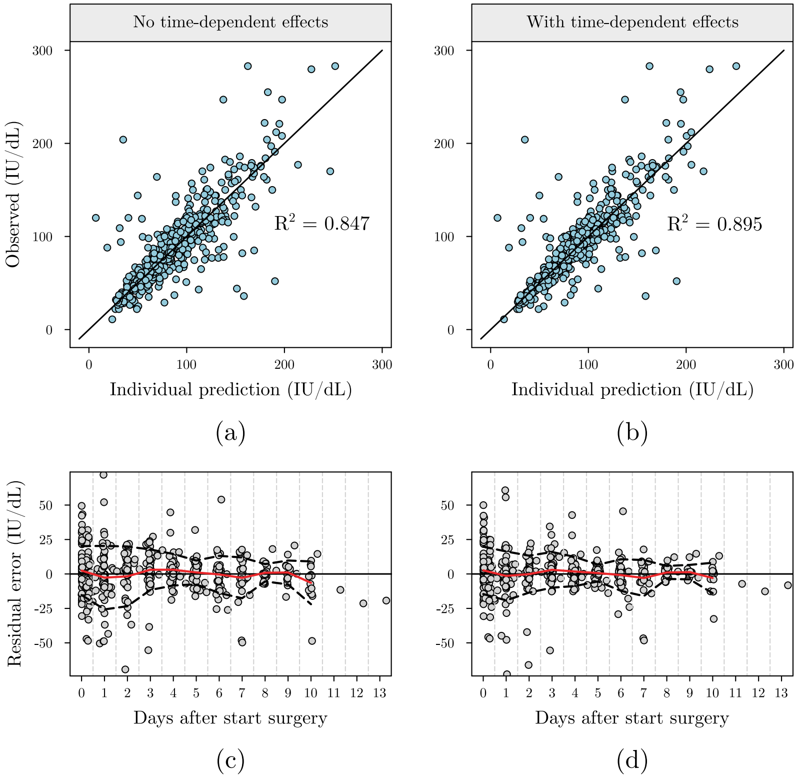


Figure 8.A.2: Prediction accuracy of the perioperative model with and without time-dependent effects on clearance. Goodness-of-fit (a & b) and Bland-Altman (c & d) plots for the perioperative models without (a & c) and with time-dependent effects (b & d). The addition of time-dependent improved the $R^2$ of model predictions, while we can also observe a lower variability of the residual error in the initial days following the surgical procedure.

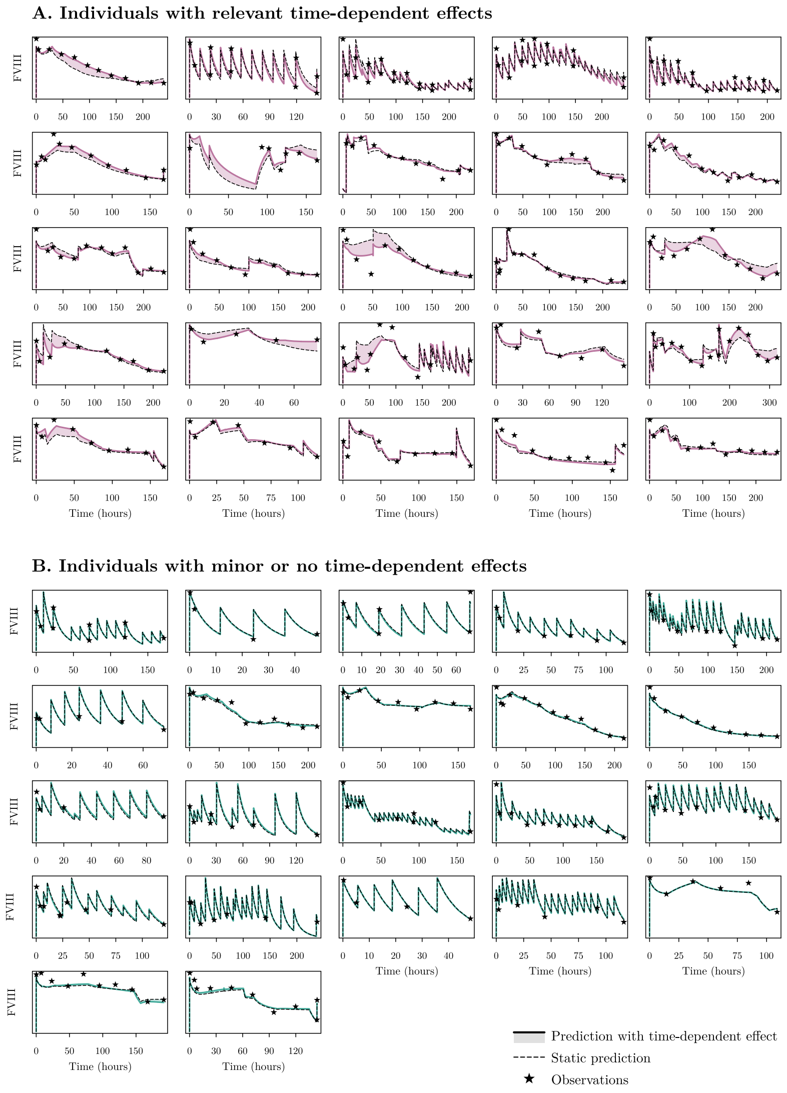


Figure 8.A.3: Overview of predictions for each of the patients with follow-up greater.

#### 8.B POPULATION PHARMACOKINETIC MODEL DEVELOPMENT

In this study, a population pharmacokinetic (PK) model for factor VIII (FVIII) was developed that can be used to transform individual PK parameters from the prophylactic setting to the perioperative setting. First, missing data was imputed using multiple imputation. We then developed a non-linear mixed effect (NLME) model on the prophylactic data. Based on individual PK parameter estimates from this model, we fitted an additional NLME model to find clinical determinants that explained how these parameters changed in the perioperative setting.

##### 8.B.1 *Missing data imputation*

Ten samples were taken from covariate distributions obtained from a recently described generative model [1]. Based on available covariates such as patient age and blood group, this generative model provides conditional distributions for the missing covariates. For example, based on patient age, we can obtain distributions over corresponding heights and body weight, allowing for the imputation of missing values for fat-free mass. A different approach was taken for the imputation of missing von Willebrand factor antigen (VWF:Ag) levels. The generative model provides distributions for which to sample from for VWF:Ag based on patient age and blood group. Instead of directly sampling from these distributions, we added a random effect component to the prophylactic population PK model that estimates the subject-specific maximum a posteriori (MAP) estimate of VWF:Ag based on the observed FVIII activity levels. We then used these MAP estimates of VWF:Ag to estimate their difference with the mean VWF:Ag measurements for each subject obtained during the perioperative setting. Perioperative VWF:Ag levels was estimated to be on average 30% higher compared to prophylactic VWF:Ag levels. For individuals missing perioperative VWF:Ag levels, missing values were imputed based on multiplying their VWF:Ag levels estimated from the prophylactic model by 1.3.

##### 8.B.2 *SHAP analysis on prophylactic data*

In order to inform the covariate selection procedure, we first performed a SHAP analysis on the prophylactic data as outlined in [2].

In a SHAP analysis, a machine learning (ML) model is fit to predict the individual PK parameter estimates obtained from a base NLME (two compartments, no covariate effects, combined error, and between-subject variability on clearance and volume of distribution) model based on the covariates. After fitting the model, a ML explanation method (SHAP) is used to identify the covariate effects learned by the ML model. In this case, we fit a Random forest model to predict the individual clearance and volume of distribution parameters based on patient body either weight & height or fat-free mass, VWF:Ag (imputed by the mode of the prior distributions from the generative model), haemophilia severity, specific FVIII concentrate administered, and the treatment centre. Since body weight / height and fat-free mass are correlated, the SHAP analysis was repeated twice, switching out body weight & height with fat-free mass. Both patient age and blood group were not evaluated, as their correlation to FVIII PK likely indirectly acts through VWF, following a causal model previously outlined in [1]. Since no between-subject variability was included on the inter-compartmental clearance and peripheral volume of distribution in the base NLME model, these parameters covariate effects could not be evaluated by the SHAP analysis. In figure A1, the resulting visualisation of covariate effects for the prophylactic data is shown.

The covariate effects on FVIII clearance (see figure 8.B.1a) and volume of distribution (figure 8.B.1b) are shown when training the model on the covariate set including fat-free mass instead of body weight and height. In addition, the effects of body weight and height on FVIII clearance (figure 8.B.1c) and volume of distribution (figure 8.B.1d) are shown when using the alternative covariate set. For the continuous covariates, we see that in the model using fat-free mass depicts more simple (linear) covariate effects compared to the model using body weight and height (figure 8.B.1c & d). In the latter model, the relationship of height to the PK parameters is non-linear, with weight having little effect. For the categorical covariates (bottom panel in figure 8.B.1a & b) we can see that patients treated in centre 2 and centre 7 have different PK parameters compared to patients from the other centres. In addition, we see differences in the PK parameters for patients receiving rFVIII-FS. Other covariates do not seem to be very important.

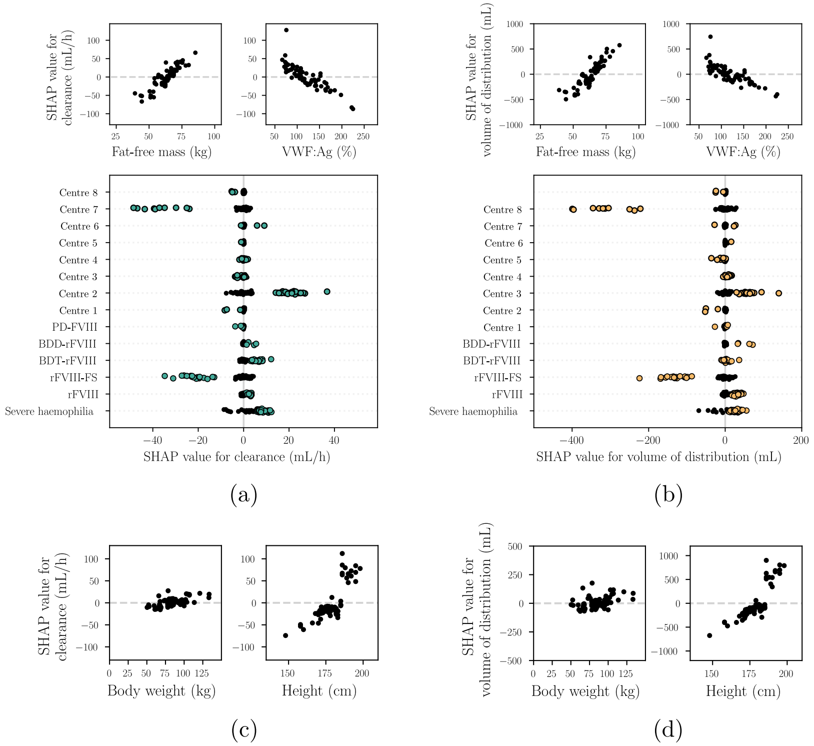


Figure 8.B.1: Visualisation of covariate effects obtained through the SHAP analysis on the prophylactic data.

##### 8.B.3 *Development of prophylactic PK model*

Based on the visualisation in figure 8.B.1, we decided to include the effect of fat-free mass on FVIII clearance and volume of distribution using a linear effect. Next, a linear effect of VWF:Ag on clearance was also added. As it is possible that there are differences in the PK between different FVIII concentrates, we added a proportional effect on all PK parameters for patients receiving rFVIII-FS. We deemed it unlikely that the specific treatment centre has a causal effect on patient PK parameters, and instead added their effect in the error model to directly affect predictions. The rationale behind this decision is that observed FVIII levels can generally be higher or lower per treatment centre, likely related to the specific measurement assay and reagents used by each of the centres. Although not indicated by the SHAP analysis, it is well known that BDD-rFVIII levels are underestimated by the one-stage assay. We thus also added a proportional effect on the predictions for patients receiving this FVIII concentrate. The initial full covariate model was thus described as follows:

$$\begin{aligned} CL &= \theta_1 \cdot \frac{ffm}{60} \cdot (1 - \theta_2 \cdot (vwf - 100\%)) \cdot \theta_3^{rFVIII-FS} \cdot \exp(\eta_1) \\ V_1 &= \theta_4 \cdot \frac{ffm}{60} \cdot \theta_5^{rFVIII-FS} \cdot \exp(\eta_2) \\ Q &= \theta_6 \cdot \theta_7^{rFVIII-FS} \\ V_2 &= \theta_8 \cdot \theta_9^{rFVIII-FS} \\ IPRED &= F \cdot \theta_{10}^{\text{centre 2}} \cdot \theta_{11}^{\text{centre 7}} \cdot \theta_{12}^{BDD-rFVIII} \end{aligned} \tag{8.1}$$

Initial parameter estimates from this model are shown in table 8.B.1.

| PARAMETER IN EQ. 8.1 | INTERPRETATION | ESTIMATE (% RSE) |
|---|---|---|
| $\theta_1$ | Typical clearance (mL/h) | 248 (6) |
| $\theta_2$ | Fold change in clearance per 100 unit increase in VWF:Ag | 0.386 (14) |
| $\theta_3$ | Fold change in clearance when patient received rFVIII-FS | 0.797 (7) |
| $\theta_4$ | Typical volume of distribution (mL) | 2630 (12) |
| $\theta_5$ | Fold change in volume of distribution when patient received rFVIII-FS | 0.867 (24) |
| $\theta_6$ | Typical inter-compartmental clearance (mL/h) | 114 (75) |
| $\theta_7$ | Fold change in inter-compartmental clearance when patient received rFVIII-FS | 2.33 (100) |
| $\theta_8$ | Typical peripheral volume of distribution (mL) | 817 (37) |
| $\theta_9$ | Fold change in typical volume of distribution when patient received rFVIII-FS | 1.09 (59) |
| $\theta_{10}$ | Fold change in prediction when patient was treated in centre 2 | 0.858 (6) |
| $\theta_{11}$ | Fold change in prediction when patient was treated in centre 7 | 1.08 (9) |
| $\theta_{12}$ | Fold change in prediction when patient received BDD-rFVIII | 0.803 (14) |
| $\omega_1$ | Standard deviation of between-subject variability on clearance | 0.184 (42) |
| $\omega_2$ | Standard deviation of between-subject variability on volume of distribution | 0.142 (95) |
| $\sigma_1$ | Standard deviation of additive error (IU/mL) | 0.0134 (14) |
| $\sigma_2$ | Standard deviation of proportional error | 0.146 (12) |

Abbreviations: RSE = relative standard error.

Table 8.B.1: Parameter estimates for the first imputation replicate of the full covariate model..

We then performed backward elimination to select covariates to be retained in the model. Each step and its result in this procedure is outlined in table 8.B.2.

After the backward elimination procedure, the final prophylactic PK model was described by the following set of equations:

| STEP | PARAMETER(S) | AVERAGE OFV | AVERAGE DOFV | DECISION |
|---|---|---|---|---|
| 1. Full covariate model | - | 977.57 | 0 | - |
| 2. Removal of rFVIII-FS from $Q$ and $V_2$ | $\theta_7$ and $\theta_9$ | 974.62 | -2.95, p >0.01 | Remove from model. |
| 3. Removal of between-subject variability on $V_1$ | $\omega_2$ | 973.54 | -1.08 (compared to 2), p >0.01 | Retained in model*. |
| 4. Removal of proportional error | $\sigma_2$ | 879.25 | -95.37 (compared to 2), p <0.01 | Retained in model. |
| 5. Removal of effect of centre 2 on $F$ | $\theta_{10}$ | 968.12 | -6.5 (compared to 2.), p >0.01 | Retained in model**. |
| 6. Removal of effect of centre 7 on $F$ | $\theta_{11}$ | 973.38 | -1.24 (compared to 2.), p >0.01 | Removed from model. |

* = Although the significance value indicated that this effect could be removed, we chose to retain it in the model. ** = significant change in the OFV for half of the imputation replicates.

Table 8.B.2: Backward elimination of model components starting from the full covariate model.

$$
\begin{aligned}
CL &= \theta_1 \cdot \frac{ffm}{60} \cdot (1 - \theta_2 \cdot (vwf - 100\%)) \cdot \theta_3^{rFVIII-FS} \cdot \exp(\eta_1) \\
V_1 &= \theta_4 \cdot \frac{ffm}{60} \cdot \theta_5^{rFVIII-FS} \cdot \exp(\eta_2) \\
Q &= \theta_6 \\
V_2 &= \theta_7 \\
IPRED &= F \cdot \theta_8^{\text{centre } 2} \cdot \theta_9^{BDD-rFVIII}
\end{aligned}
\tag{8.2}
$$

An overview of the final model parameters are shown in table 8.B.3.

##### 8.B.4 *SHAP analysis on perioperative data*

After fitting the prophylactic model, we collected individual estimates of the PK parameters for each of the subjects. We then fitted a base NLME model to the perioperative data. We then performed a SHAP analysis on the difference between the perioperative PK parameters from the base NLME model and the individual PK parameters obtained from the prophylactic setting. This way the SHAP analysis

| PARAMETER IN EQ. 8.2 | INTERPRETATION | ESTIMATE (% RSE) |
|---|---|---|
| $\theta_1$ | Typical clearance (mL/h) | 246 (12) |
| $\theta_2$ | Change in clearance per 100 unit increase in VWF:Ag | 0.387 (4) |
| $\theta_3$ | Change in clearance when patient received rFVIII-FS | 0.79 (8) |
| $\theta_4$ | Typical volume of distribution (mL) | 2560 (6) |
| $\theta_5$ | Change in typical volume of distribution when patient received rFVIII-FS | 0.941 (7) |
| $\theta_6$ | Typical inter-compartmental clearance (mL/h) | 132 (30) |
| $\theta_7$ | Typical peripheral volume of distribution (mL) | 800 (16) |
| $\theta_8$ | Change in prediction when patient was treated in centre 2 | 0.853 (8) |
| $\theta_9$ | Change in prediction when patient received BDD-rFVIII | 0.794 (9) |
| $\omega_1$ | Standard deviation of between-subject variability on clearance | 0.183 (42) |
| $\omega_2$ | Standard deviation of between-subject variability on volume of distribution | 0.121 (21) |
| $\sigma_1$ | Standard deviation of additive error (IU/mL) | 0.013 (19) |
| $\sigma_2$ | Standard deviation of proportional error | 0.155 (25) |

Abbreviations: RSE = relative standard error.

Table 8.B.3: Parameter estimates for the first imputation replicate of the final prophylactic model.

identifies clinical markers important to explain changes in PK during the perioperative setting. The resulting visualisations are shown in figure A2.

#### 8.B.5 *Perioperative PK model*

For the perioperative population PK model, covariate analysis is complicated by relevant correlations between the covariates (see figure 8.3.2 in the main manuscript). Based on the correlation matrix from figure 8.A.1 and the SHAP analysis in figure 8.B.2, we decided on including covariates related to the intensity / risk of the surgical procedure. This included pre-assessed surgical risk score, blood loss during and after surgery, surgery duration, administration of NaCl, and the change in VWF:Ag levels compared to the prophylactic setting. Again, a full model approach was taken. The initial full model was represented by the following set of equations:

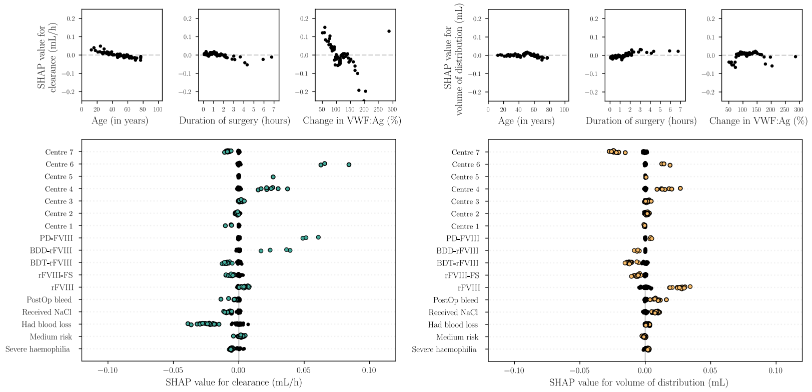


(a) (b)

Figure 8.B.2: SHAP analysis for the change in FVIII PK during the perioperative setting.
The covariate effects on the change FVIII clearance (a) and volume of distribution (b) in the perioperative setting compared to the prophylactic setting are shown For the continuous covariates, we see that VWF:Ag has an effect on the change in perioperative FVIII clearance. Furthermore, we notice lower clearance for patients with blood loss during surgery, and higher clearance in patients treated in Nijmegen. For volume of distribution, we notice potential effect for patients treated in centre 7 and those receiving rFVIII.

$$
\begin{aligned}
CL &= CL^{p} \cdot \theta_1 \cdot \theta_2^{PostBL} \cdot \theta_3^{BL} \cdot \theta_4^{DUR>2h} \cdot \theta_5^{VWFdecrease} \cdot \theta_6^{VWF+40\%} \cdot \exp(\eta_1) \\
V_1 &= V_1^{p} \cdot \theta_7 \cdot \theta_8^{Bleeding} \cdot \exp(\eta_2) \\
Q &= Q^{p} \cdot \theta_9 \\
V_2 &= V_2^{p} \cdot \theta_{10} \\
IPRED &= F \cdot \theta_{11}^{\text{centre 2}} \cdot \theta_{12}^{\text{centre 7}} \cdot \theta_{13}^{BDD-rFVIII}
\end{aligned}
\tag{8.3}
$$

Again, we performed backward elimination to select covariates to be retained in the model. Each step and its result in this procedure is outlined in table 8.B.5.

After the backward elimination procedure, the final perioperative PK model was described by the following set of equations:

| PARAMETER IN EQ. 8.3 | INTERPRETATION | ESTIMATE (% RSE) |
|---|---|---|
| $\theta_1$ | Change in clearance with respect to the prophylactic setting. | 0.92 (6) |
| $\theta_2$ | Change in clearance when a post-operative bleed occurred. | 0.841 (7) |
| $\theta_3$ | Change in clearance when perioperative bleeding occurred. | 1.02 (7) |
| $\theta_4$ | Change in clearance when the duration of surgery was greater than two hours. | 0.853 (7) |
| $\theta_5$ | Change in clearance when VWF:Ag levels were lower than in the prophylactic setting | 1.14 (6) |
| $\theta_6$ | Change in clearance when VWF:Ag levels increased by more than 40%. | 0.902 (8) |
| $\theta_7$ | Change in volume of distribution with respect to the prophylactic setting. | 1.15 (4) |
| $\theta_8$ | Change in volume of distribution when a bleed occurred. | 1.13 (5) |
| $\theta_9$ | Change in inter-compartmental clearance with respect to the prophylactic setting. | 0.952 (20) |
| $\theta_{10}$ | Change in peripheral volume of distribution with respect to the prophylactic setting. | 1.79 (12) |
| $\theta_{11}$ | Change in prediction when patient was treated in centre 2 | 1.04 (4) |
| $\theta_{12}$ | Change in prediction when patient was treated in centre 7 | 1.09 (5) |
| $\theta_{13}$ | Change in prediction when patient received BDD-rFVIII | 0.814 (7) |
| $\omega_1$ | Standard deviation of between-subject variability on clearance | 0.174 (23) |
| $\omega_2$ | Standard deviation of between-subject variability on volume of distribution | 0.101 (53) |
| $\sigma_1$ | Standard deviation of additive error (IU/mL) | 0.013 (Fixed) |
| $\sigma_2$ | Standard deviation of proportional error | 0.179 (4) |

Abbreviations: RSE = relative standard error.

Table 8.B.4: Parameter estimates for the first imputation replicate of the full covariate model fit to the perioperative data.

$$\begin{aligned}
CL &= CL^p \cdot \theta_1 \cdot \theta_2^{PostOpBleed} \cdot \theta_3^{Duration>2h} \cdot \theta_4^{VWFdecrease} \cdot \exp(\eta_1) \\
V_1 &= V_1^p \cdot \theta_5 \cdot \theta_6^{Bleeding} \cdot \exp(\eta_2) \\
Q &= Q^p \cdot \theta_7 \\
V_2 &= V_2^p \cdot \theta_8 \\
IPRED &= F \cdot \theta_9^{BDD-rFVIII}
\end{aligned} \tag{8.4}$$

An overview of the final model parameters are shown in table 8.B.6.

| STEP | PARAMETER | AVERAGE OFV | AVERAGE DOFV | DECISION |
|---|---|---|---|---|
| 1. Full covariate model | - | -1325.76 | 0 | - |
| 2. Remove effect perioperative bleeding on clearance | $\theta_3$ | -1325.68 | +0.084 (compared to 1), p >0.01 | Removed from model. |
| 3. Remove effect centre 2 on the prediction | $\theta_{11}$ | -1324.47 | +1.21 (compared to 2), p >0.01 | Removed from model. |
| 4. Remove effect centre 7 on the prediction | $\theta_{12}$ | -1321.79 | +2.68 (compared to 3), p >0.01 | Removed from model. |
| 5. Remove effect of surgery on peripheral clearance | $\theta_9$ | -1321.34 | +0.45 (compared to 4), p >0.01 | Removed from model. |
| 6. Replacing surgery duration with surgical risk score. | $\theta_4$ | -1315.97 | +5.37 (compared to 4), p >0.01 | Retained in model. |
| 7. Removing effect increase of VWF on clearance | $\theta_6$ | -1319.97 | +1.37 (compared to 5), p >0.01 | Removed from model. |

Abbreviations: RSE = relative standard error.

Table 8.B.5: Backward elimination of model components starting from the full covariate model.

#### 8.C IMPLEMENTATION OF TIME-DEPENDENT EFFECTS

We used Gaussian Processes in order to identify time-dependent effects on FVIII clearance during the perioperative setting. Gaussian Processes are an extension of the multivariate normal distribution in infinite dimensions [1]. At each time point, a Gaussian Process outputs a Gaussian (Normal) distribution. In typical non-linear mixed effects models, the random effect is described by a single (multivariate) Normal distribution, and a single estimate of its effects is given for the entire time frame. This can be extended to inter-occasion variability, where we allow re-estimation of the random effect after a certain period of time. Simply estimating a new Gaussian distribution at each time point is prone to overfit the data. Gaussian Processes can reduce overfitting through the use of a kernel function. The kernel function

| PARAMETER IN EQ. 8.4 | INTERPRETATION | ESTIMATE (% RSE) |
|---|---|---|
| $\theta_1$ | Change in clearance with respect to the prophylactic setting. | 0.875 (4) |
| $\theta_2$ | Change in clearance when a post-operative bleed occurred. | 0.844 (10) |
| $\theta_3$ | Change in clearance when the duration of surgery was greater than two hours. | 0.845 (6) |
| $\theta_4$ | Change in clearance when VWF:Ag levels were lower than in the prophylactic setting | 1.16 (5) |
| $\theta_5$ | Change in volume of distribution with respect to the prophylactic setting. | 1.11 (2) |
| $\theta_6$ | Change in volume of distribution when a bleed occurred. | 1.15 (4) |
| $\theta_7$ | Change in peripheral volume of distribution with respect to the prophylactic setting. | 1.73 (18) |
| $\theta_8$ | Change in prediction when patient received BDD-rFVIII | 0.792 (12) |
| $\omega_1$ | Standard deviation of between-subject variability on clearance | 0.176 (24) |
| $\omega_2$ | Standard deviation of between-subject variability on volume of distribution | 0.109 (51) |
| $\sigma_1$ | Standard deviation of additive error (IU/mL) | 0.013 (Fixed) |
| $\sigma_2$ | Standard deviation of proportional error | 0.179 (7) |

Abbreviations: RSE = relative standard error.

Table 8.B.6: Parameter estimates for the first imputation replicate of the final perioperative model..

describes the correlation between each Gaussian variable over time. When correlation is high, distant variables (in time) are more similar. When correlation is low, the Gaussian variables appear to be more randomly distributed. Gaussian Processes are also often framed as representing a distribution over functions, with the kernel function describing the complexity of the functions.

The choice of kernel function can be used to add prior knowledge to model structure, improving performance on smaller data sets. We used a Matérn32 kernel, which results in somewhat smooth functions. The kernel has two parameters: the variance, and the length scale. The variance represents the variability in the means for each of the Gaussian variables. A higher variance means that the means of the Gaussian variables can take on larger values. In our use-case, higher variances reflect larger time-dependent effects. Next, the length scale determines the correlation between the random variables over time. Smaller length scales mean that the time-dependent effect can change more quickly in time. If the length scale is too small, the model becomes more prone to overfit the data. These parameters are referenced as the kernel hyper-parameters.

In order to learn these parameters from the data we performed Variational Expectation Maximisation. We first fit a Gaussian Process to the data from each specific subject, and then optimise the hyper-parameters based on all subject-specific models. Since the estimation of random effects in non-linear mixed effect models is not exact, we need to fit approximate Gaussian Processes to obtain posterior distributions over the time-dependent effects. For this purpose we use Sparse Variational Gaussian Processes (SVGPs) [2]. Gaussian Processes are normally directly fitted based on observed data. Since the random (time-dependent) effects are not observed, we instead optimise the SVGP based on pseudo-points using Variational Inference. This means that we learn inputs that influence the shape of the SVGP in such a way that the data log likelihood is maximised. Pseudo-inputs were estimated at each 6 hour interval. After obtaining each subject-specific SVGP model, the SVGP parameters were fixed and the model hyper-parameters were optimised. Alongside the kernel parameters, we also optimised the proportional error estimated by the perioperative population PK model. This estimate was slightly higher than in the prophylactic setting (0.179 vs. 0.155), which is likely due to the presence of time-dependent effects resulting in an over-estimation of the error. Additive error was fixed to 1.3 IU/dL. Initial estimates for the kernel hyper-parameters were a variance of 0.1 and a length scale of 1/30. Initial estimates were selected based on the shape of samples from the resulting Gaussian Process prior. Final hyper-parameter estimates were a variance of 0.01 and a length scale of 1/93. Re-estimated proportional error was 0.151, similar to the estimate obtained in the prophylactic population PK model (0.155). A comparison between the Gaussian Process prior and the model with optimised hyper-parameters is shown in figure B1.

We see that the lower variance estimated for the optimised model results smaller maximal changes in FVIII clearance, while the lower length scale also results in more smooth functions.

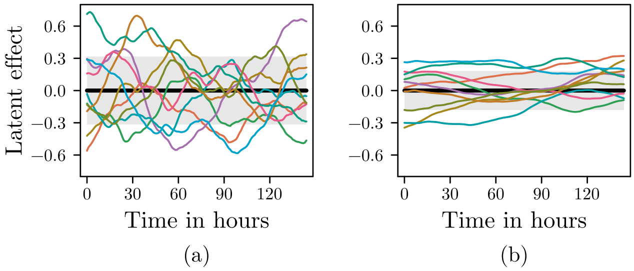


Figure 8.C.1: Comparison between samples from the Gaussian Process prior and optimised model. Here we see that samples (coloured lines) from the untrained prior (a; black line and filled area) resulted in both stronger and less smooth effects over time compared to the final optimised model (b).
